# Altered Brain Glucose Metabolism in COVID-19 disease: An activation likelihood estimation Meta-analysis

**DOI:** 10.1101/2024.04.30.24306508

**Authors:** Dongju Kang, Hyunji Jung, Kyoungjune Pak

## Abstract

**Purpose:** COVID-19, caused by the SARS-CoV-2 virus, has significantly altered modern society and lifestyles. We investigated its impact on brain glucose metabolism by meta-analyzing existing studies that utilized 18F-fluorodeoxyglucose (FDG) positron emission tomography (PET) scans of the brain.

**Methods:** We conducted a systematic search of MEDLINE and EMBASE databases from inception to August 2023 for English-language publications using the keywords “positron emission tomography,” “single-photon emission computed tomography,” and “COVID-19.” We included original research articles that reported changes in brain glucose metabolism following COVID-19 infection. ALE values from these studies were aggregated and tested against a null hypothesis that anticipated a random distribution of ALE values, which proved to be significantly higher than chance.

**Results:** We identified eight papers that met our inclusion criteria. Significant increases in brain glucose metabolism were noted in the left anterior cingulate gyrus, right thalamus, and brainstem. In children with COVID-19, decreased glucose metabolism was observed in the right and left cerebellum, left amygdala/hippocampus, left anterior cingulate gyrus, and right amygdala. In adults with COVID-19, decreased metabolism was seen in the right temporal lobe, brainstem (acute phase), left posterior cingulate gyrus, left precuneus, right cerebellum, right insula, right anterior cingulate gyrus, left occipital lobe, and left globus pallidus (chronic phase).

**Conclusion:** COVID-19 impacts brain glucose metabolism, typically manifesting as areas of decreased metabolism in ^18^F-FDG PET scans, though increases are also observed. These changes in metabolism vary with the patient’s age and the time elapsed between the diagnosis of COVID-19 and the PET scan.

## INTRODUCTION

COVID-19, caused by the SARS-CoV-2 virus, has reshaped societal norms and presented significant public health challenges worldwide. Its rapid spread prompted global interventions, including social distancing and mask mandates, to curb transmission. By April 2024, the disease had resulted in over 700 million cases and 7 million deaths worldwide ^1^. The clinical spectrum of COVID-19 ranges from common symptoms like fever and cough ^2^ to severe complications affecting various organ systems, including the nervous system, circulatory system, and kidneys ^3^. Neurological complications are not rare among COVID-19 patients, with disruptions in olfactory and gustatory functions being especially prevalent. In severe instances, patients may suffer from seizures and altered consciousness ^4^. These neurological complications often persist well beyond the acute phase of the illness and can be studied using neuroimaging techniques.

Various brain imaging modalities have been utilized to examine changes in patients with COVID-19. Brain magnetic resonance imaging (MRI) has revealed extensive confluent or multifocal white matter lesions, microhemorrhages, diffusion restriction, and enhancement in COVID-19 patients^5^. Additionally, multiple white matter lesions have been documented^6^. Beyond structural changes, blood oxygen level-dependent functional MRI (BOLD-fMRI) has demonstrated enhanced signals in the right piriform cortex and right anterior cingulate gyrus^7^. Given that the human brain primarily uses glucose as its energy source^8, 9, 18^F-FDG PET scans were performed to assess neuronal activity by comparing brain glucose metabolism in COVID-19 patients with that of healthy controls. Two studies reported increased brain glucose metabolism in COVID-19 patients ^10, 11^ while others noted a decrease^10-17^. Consequently, it is pertinent to meta-analyze previous studies on changes in brain glucose metabolism in COVID-19. In this study, we categorized the existing publications into two groups based on the metabolic changes: 1) hypermetabolism and 2) hypometabolism in COVID-19 patients. We then conducted a meta-analysis on studies of hypometabolism, focusing on 1) the patient’s age and 2) the time interval between the diagnosis of COVID-19 and the PET scan.

## MATERIALS AND METHODS

### Data Search, Study Selection, and Data Extraction

We conducted a systematic search of MEDLINE and EMBASE databases, from their inception to August 2023, for English-language publications using the keywords “positron emission tomography”, “single-photon emission computed tomography”, and “COVID-19.” The inclusion criteria were limited to original research articles that reported changes in brain glucose metabolism following COVID-19 infection. The results were reported as coordinates in a normalized standard stereotactic space (either Talairach or Montreal Neurological Institute space). Studies that focused solely on regions of interest were excluded. Data were independently extracted by two reviewers from the publications, and the following information was recorded: year of publication, country of affiliation of the corresponding authors, name of the journal, number of studies included, database search, radiopharmaceuticals used, coordinates, and number of subjects included.

### Meta-analysis Algorithm

The software GingerALE ver 3.0.2 (Research Imaging Institute, University of Texas Health Science Center at San Antonio, TX, USA) was used to transform all reported coordinates into stereotactic Montreal Neurological Institute space. The method employed in this study was a variation of the original ALE by Turkeltaub et al. ^18^, which was later modified by Eickhoff et al. ^19, 20^. For each experiment, the modeled activation map was calculated by determining the maximum value across each Gaussian focus. The width of the Gaussian probability distribution was determined individually for each experiment based on empirical estimates of between-subject variability derived from the number of subjects in each study. For each voxel, the ALE value was calculated from the union of the modeled activation maps. ALE values were combined across studies and tested against a null hypothesis of a random distribution of ALE values, which were higher than expected by chance. A threshold of FWE 0.05 was applied for both cluster-level forming thresholds and for cluster-level inference on the resulting ALE map.

## RESULTS

### Literature Search and Study Characteristics

A total of 1,078 records were identified from electronic (n=1,075) and manual (n=3) searches. Of these, 417 were excluded prior to screening. To assess the eligibility of the remaining 661 studies, we screened them based on their titles and abstracts. A total of 44 papers were selected for full-text review to further evaluate their suitability. Consequently, eight papers were deemed suitable for inclusion in the study ^10-17^. The detailed procedure is illustrated in Figure 1, and the characteristics of the studies are summarized in Table 1.

**Figure 1.**
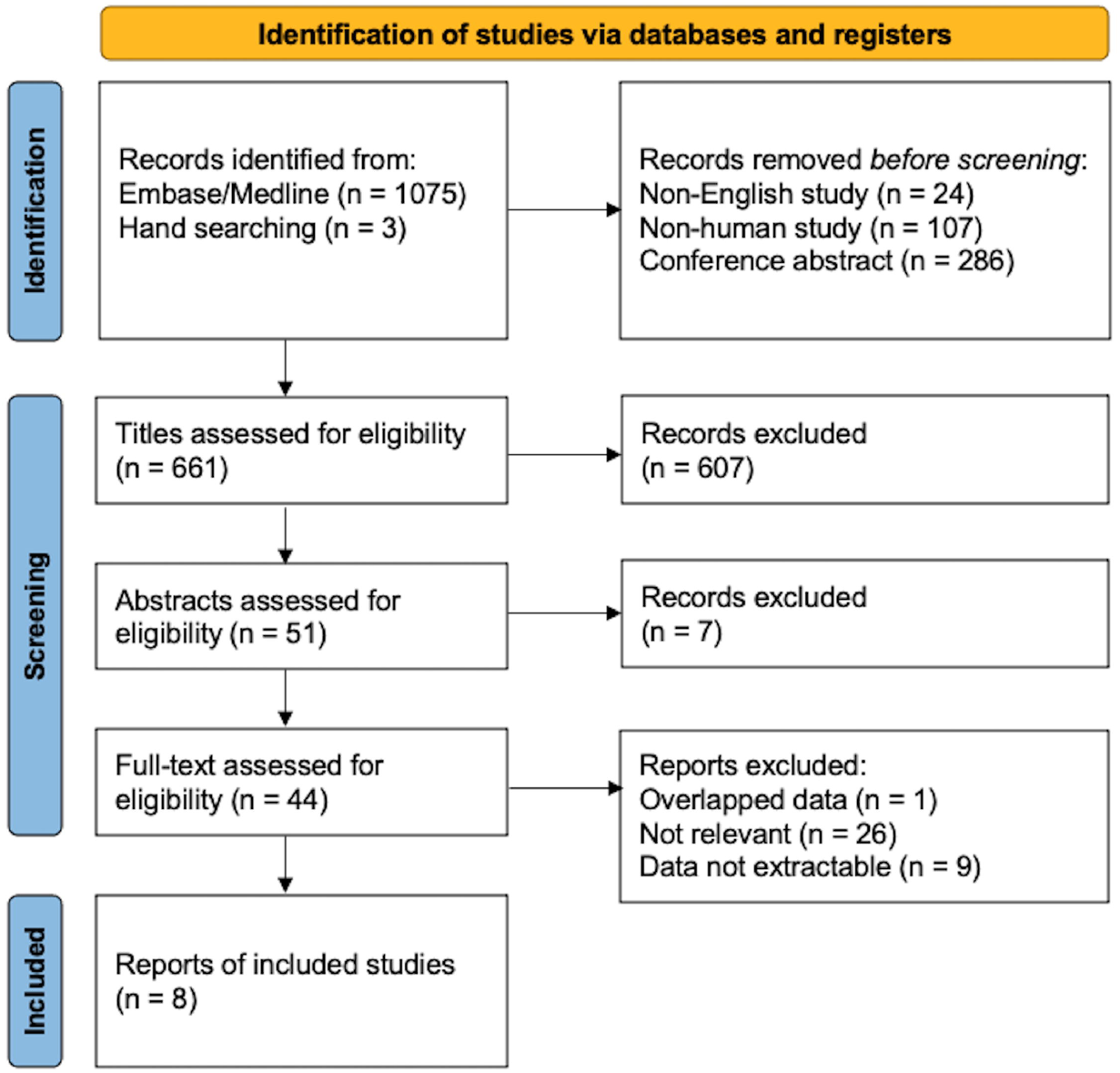
Flowchart for the identification of eligible studies

### Anatomical Likelihood Estimate analysis

#### A. Increased Brain Glucose Metabolism in COVID-19 disease

Two studies reported increased brain glucose metabolism in 19 COVID-19 patients compared to 58 controls ^10, 11^. A significant increase in brain glucose metabolism was observed in the left anterior cingulate gyrus (cluster size:13,176 mm^3^, x -30; y 38; z 0, maximum ALE value of 0.0095), right thalamus (cluster size:13,176 mm^3^, x 24; y -18; z 8, maximum ALE value of 0.0095), and brainstem (cluster size:11,704 mm^3^, x 12; y -25; z -37, maximum ALE value of 0.0089) (Figure 2).

**Figure 2.**
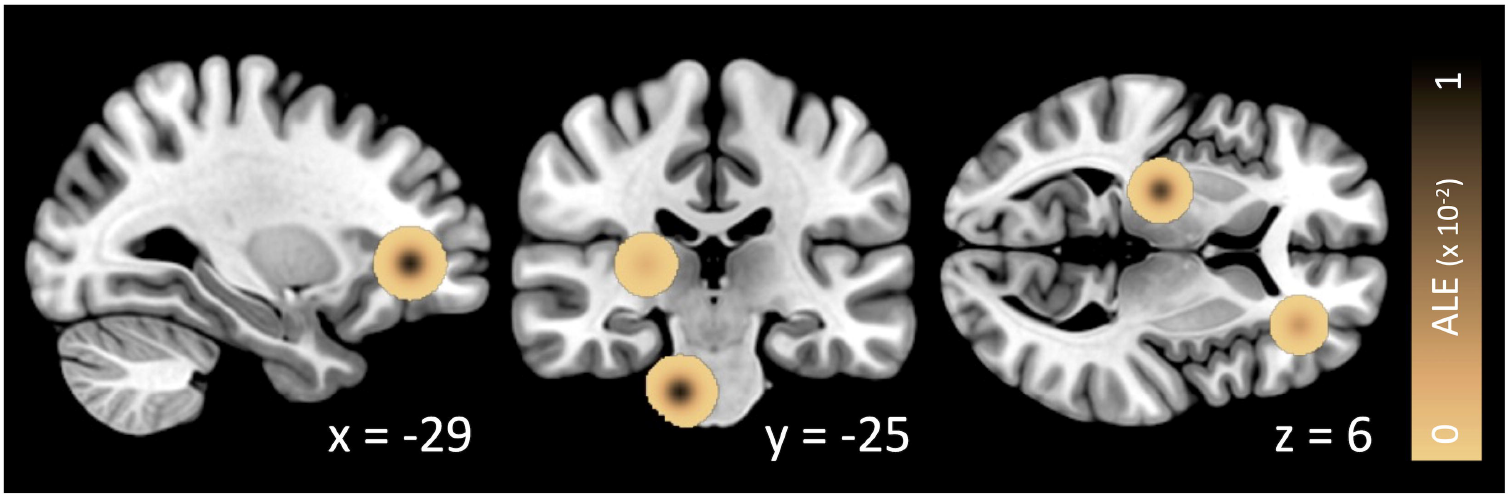
ALE analysis of increased brain glucose metabolism in COVID-19 disease

#### B. Decreased Brain Glucose Metabolism in COVID-19 disease

Studies on hypometabolism in COVID-19 were categorized into subgroups according to 1) patient age and 2) the time interval between COVID-19 diagnosis and PET scanning.

##### B1. COVID-19 disease in Children and Adults

We analyzed eight studies, comprising six studies on adults ^10-15^, and two studies on children^16, 17^. Children were defined as those aged < 18 years old. In children with COVID-19 disease, decreased glucose metabolism was observed in five regions, including the right cerebellum (cluster size:35,928 mm^3^, x 20; y -70; z -42, maximum ALE value of 0.0089), left cerebellum (cluster size:18,960 mm^3^, x -30; y -52; z -44, maximum ALE value of 0.0088), left amygdala/hippocampus (cluster size:16,224 mm^3^, x -26; y -4; z -24, maximum ALE value of 0.0088), left anterior cingulate gyrus (cluster size:9,896 mm^3^, x -18; y 46; z -4, maximum ALE value of 0.0086), right amygdala (cluster size:9,536 mm^3^, x 36; y 0; z -28, maximum ALE value of 0.0088). In adults with COVID-19, one hypometabolic cluster was found in the right temporal lobe (cluster size:13,648 mm^3^, × 50; y -24; z -18, maximum ALE value of 0.0173) (Figure 3).

**Figure 3.**
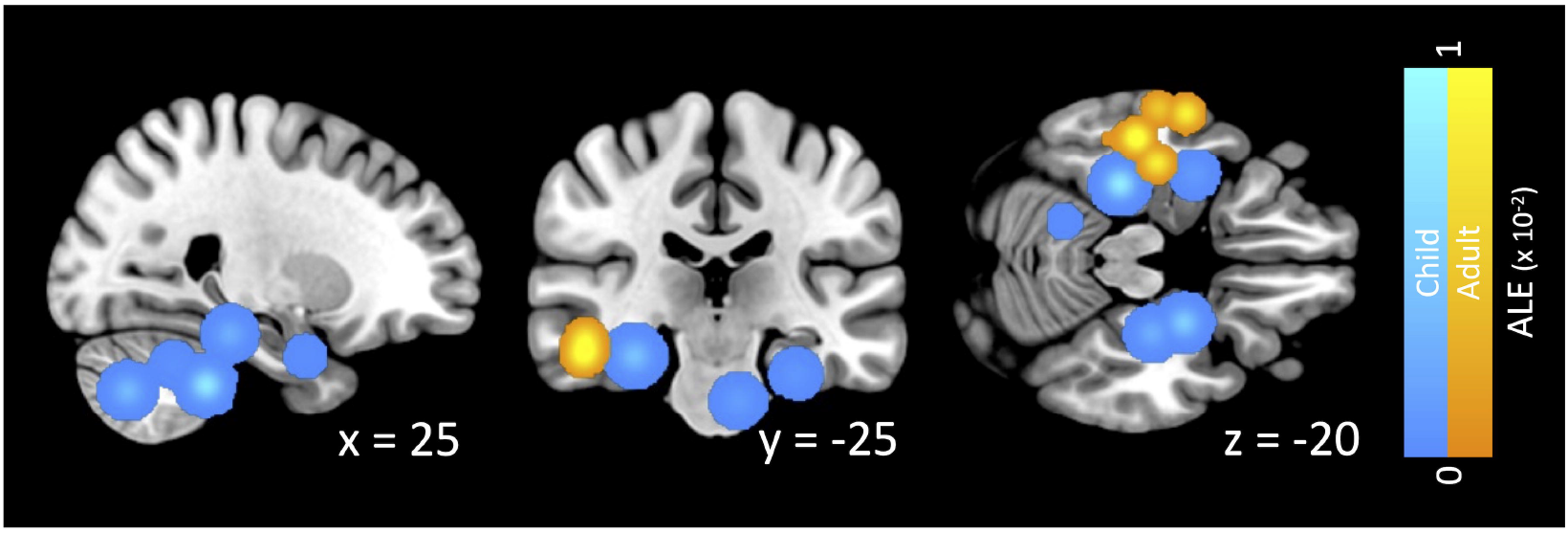
ALE analysis of decreased brain glucose metabolism in COVID-19 disease (blue: child, orange: adult)

##### B2. COVID-19 disease in acute and chronic phases

We categorized the studies into two groups based on the time interval between COVID-19 diagnosis and PET scanning. PET scans conducted within three months of infection were classified as the acute phase, while those performed after three months were considered the chronic phase.

In the acute phase of COVID-19, hypometabolism was observed in the right temporal lobe (cluster size:26,344 mm^3^, x 50; y -24; z -18, maximum ALE value of 0.0173) and brainstem (cluster size:11,200 mm^3^, x 8; y -22; z -24, maximum ALE value of 0.010). In the chronic phase of COVID-19 disease, hypometabolism was observed in 7 clusters: left posterior cingulate gyrus (cluster size: 33,728 mm^3^, x -6; y -70.3; z 12, maximum ALE value of 0.0030), left precuneus (cluster size: 15,784 mm^3^, x -34; y -78; z 40.3, maximum ALE value of 0.0010), right cerebellum (cluster size: 6,776 mm^3^, x 42; y -50; z -46, maximum ALE value of 0.0096), right insula (cluster size:6,776 mm^3^, x 46; y 14; z -2, maximum ALE value of 0.0096), right anterior cingulate gyrus (cluster size:6,560 mm^3^, x 4; y 13; z -16, maximum ALE value of 0.0092), left occipital lobe (cluster size:6,560 mm^3^, x -18; y -46; z -3, maximum ALE value of 0.0097), left globus pallidus (cluster size:6,392 mm^3^, x -22; y -12; z 4, maximum ALE value of 0.0100) (Figure 4).

**Figure 4.**
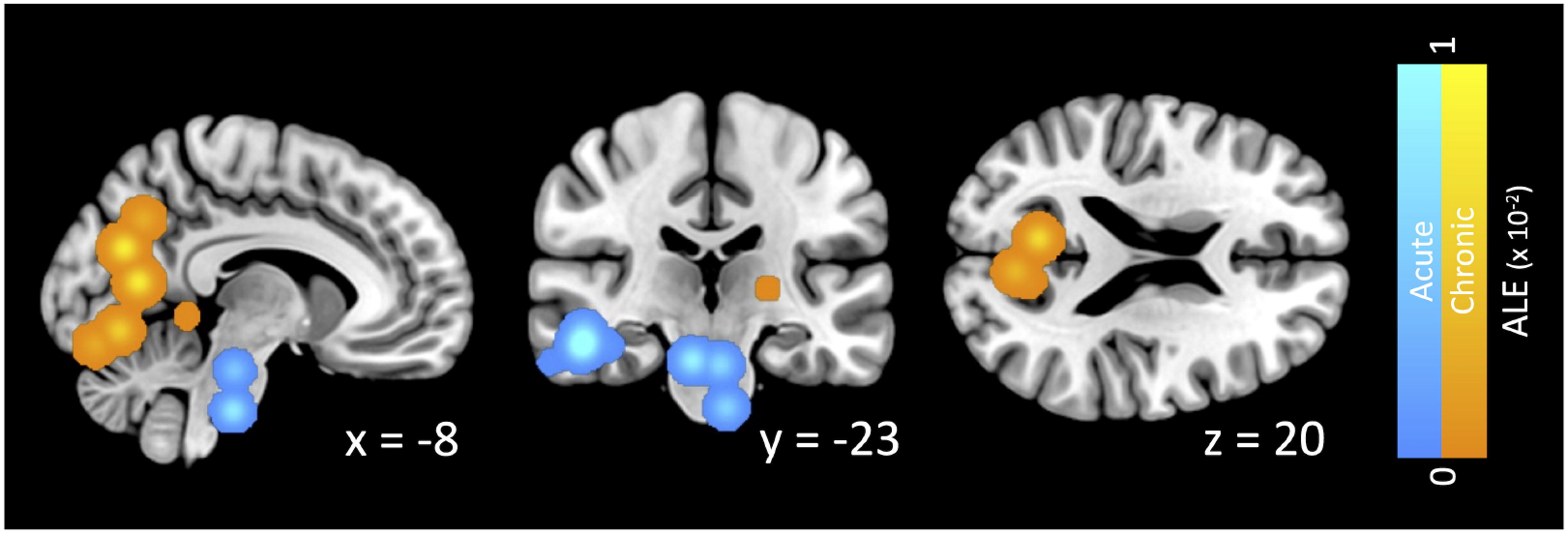
ALE analysis of decreased brain glucose metabolism of adults in COVID-19 disease (blue: acute disease, orange: chronic disease)

## DISCUSSION

COVID-19 has altered not only our lifestyle but also our brain glucose metabolism. It typically appears on ^18^F-FDG PET scans as areas of decreased brain glucose metabolism, although there may also be areas of increased metabolism. Decreases in brain glucose metabolism have been observed in the cerebellum, left amygdala/hippocampus, cingulate gyrus, right amygdala, right insula, and right temporal lobe. This reduction in metabolism varies depending on the patient’s age and the time elapsed between the COVID-19 diagnosis and the PET scan. Conversely, increased brain glucose metabolism has been noted in the left anterior cingulate gyrus, right thalamus, and brainstem.

COVID-19, caused by the SARS-CoV-2 virus, has significantly altered modern society and lifestyles. The first human infection was reported in Wuhan, China, in November 2019, and on March 11, 2020, the World Health Organization (WHO) declared the outbreak a global pandemic^21^. The virus can be transmitted through small liquid particles, which come from when infected individuals cough, sneeze, talk, sing, or breathe ^21^. Due to the rapid transmission rate in the early stages, many countries adopted social distancing and mask mandates to prevent the collapse of healthcare systems and slow the spread ^22^. As of April 2024, there have been over 700 million reported cases and over 7 million deaths^1^. COVID-19 presents with various symptoms, including fever, fatigue, dry cough, anorexia, and dyspnea ^23^. A common in-hospital complication among critically ill patients is barotrauma, which can manifest as pneumothorax, pneumomediastinum, or subcutaneous emphysema ^3^. The virus is also associated with neurological symptoms such as dizziness, headaches, disturbances in olfactory function, impaired consciousness, and seizures^4^. Among these, disturbance in olfactory function was the most prevalent, with a reported prevalence of 31.4%, followed by taste disturbance at 28.1% ^24^. More severe cases, such as seizures, have a prevalence of 1.1% ^24^. These neurological complications can be severe, especially in critically ill patients ^25^. In patients with Acute Respiratory Distress Syndrome attributed to COVID-19 disease, neurological findings were documented in nearly 80% of cases ^26^. Non-negligible rates of neurological complications persist for months or years ^27^. Moreover, COVID-19 can change brain structures for a longer period ^28^. Therefore, we investigated how COVID-19 affects brain glucose metabolism by meta-analyzing studies on brain ^18^F-FDG PET scans.

The human brain primarily uses glucose as its main energy source; therefore, brain glucose metabolism, as assessed by ^18^F-FDG PET, can be used to quantify neuronal activity^8, 9^. ^18^F-FDG PET plays a crucial role in diagnosing neurodegenerative disorders. For example, Alzheimer’s disease typically shows a distinct pattern of decreased glucose metabolism in the posterior cingulate cortex, precuneus, and temporoparietal cortex^29. 18^F-FDG PET is also effective in diagnosing and evaluating neurological disorders in pediatric patients. In cases of pediatric epilepsy, ^18^F-FDG PET helps in identifying the focal epileptogenic zone, thus enabling more precise surgical interventions ^30^. By comparing the brain glucose metabolism of patients with COVID-19 to that of healthy controls, we identified the brain regions affected by COVID-19.

Various brain imaging modalities have been employed to investigate changes in COVID-19 patients. Brain MRI has revealed extensive confluent or multifocal white matter lesions, microhemorrhages, diffusion restriction, and enhancement in these patients ^5^. More than 70% of patients exhibited multiple white matter lesions on brain MRI scans^6^. The application of Blood Oxygen Level Dependent-functional MRI indicated enhanced signals in the right piriform cortex and right anterior cingulate gyrus ^7^. Additionally, changes in brain volume have been reported. The bilateral olfactory cortex, hippocampus, insula, left Heschl’s gyrus, left Rolandic operculum, and right cingulate gyrus showed an increase in brain volume^31^, whereas an overall decrease in white matter was reported in COVID-19 ^32^. Furthermore, an increase in cortical thickness has been observed ^33^.

These diverse findings highlight the multifaceted impact of COVID-19 on the brain and underscore the importance of comprehensive neuroimaging approaches to uncover the complexities associated with post-infection neurological changes.

In this study, hypermetabolism was observed in the left anterior cingulate gyrus, right thalamus, and brainstem. The anterior cingulate gyrus is involved in the dynamic control of attention and efficient response selection^34^, the thalamus is a relay center of both sensory and motor mechanisms ^35^, and the brainstem is involved in respiratory, cardiovascular, gastrointestinal, and neurological functions ^36^. SARS-CoV-2 can cross the blood-brain barrier, triggering inflammation that leads to increased brain glucose metabolism on ^18^F-FDG PET scans. Therefore, these areas of hypermetabolism may be associated with symptoms such as fatigue, brain fog, sensorimotor issues, and sleep disturbances in COVID-19^37^. Hypometabolism was observed in the cerebellum, left amygdala/hippocampus, cingulate gyrus, right amygdala, right insula, and right temporal lobe. We found that hypometabolism in COVID-19 depends on the patient’s age and the time interval between the COVID-19 diagnosis and PET scanning. In children, hypometabolism was primarily observed in the cerebellum, which is involved in motor control, sensory perception, and higher-level cognitive processes, potentially explaining the decreased functioning in these domains post-COVID infection ^38^.

In adults, hypometabolism has been noted in the right temporal lobe, right parahippocampal gyrus, and right hippocampus, areas responsible for declarative memory, language production, cognition, and creative insight. This may contribute to common COVID-19 symptoms such as brain fog, memory problems, and fatigue.

This study had several limitations. First, it was based on a limited number of studies. Additionally, the overall number of participants included in this analysis was relatively small. Moreover, the inclusion criteria were restricted to studies that presented findings in the form of coordinates within a standardized stereotactic space, while studies utilizing region-of-interest analysis were excluded. This selection criterion could potentially impact the study outcomes. Finally, COVID-19 is a relatively new disease, and its long-term effects on brain glucose metabolism have not yet been thoroughly investigated.

In conclusion, COVID-19 alters glucose metabolism in the brain. It typically appears on ^18^F-FDG PET scans as areas of decreased brain glucose metabolism; however, there may also be areas of increased metabolism. This variation in brain glucose metabolism is dependent on the patient’s age and the time interval between the COVID-19 diagnosis and PET scanning.

## Data Availability

All data produced in the present study are available upon reasonable request to the authors

