## Supplementary material for "Altered Brain Glucose Metabolism in COVID-19 disease: An activation likelihood estimation Meta-analysis": table

**TABLES**

Table 1. Studies included in the meta-analysis

| Studies | Year | Country | No. | | Time interval between COVID-19 disease and PET scanning (m) | Coordinate |
| --- | --- | --- | --- | --- | --- | --- |
|  |  |  | Patients | Controls |  |  |
| Debs P. et al. | 2023 | USA | 45 | 52 | 6.57 | MNI |
| Goehringer F. et al. | 2023 | France | 28 | 28 | 16.4 | MNI |
| Cocciolillo F. et al. | 2022 | Italy | 3 | 19 | 4.83 | MNI |
| Guedj E. et al. | 2021 | France | 35 | 44 | 3.19 | Talairach |
| Kas A. et al. | 2021 | France | 7 | 32 | Acute | MNI |
|  |  |  |  |  | 1 |  |
|  |  |  |  |  | 6 |  |
| Niesen M. et al. | 2021 | Belgium | 12 | 26 | 0.5 | MNI |
| Donegani MI. et al. | 2021 | Italy | 14 | 61 | 1-3 | Talairach |
| Morand A. et al. | 2021 | France | 7 | 21 | ≥ 6 | Talairach |
